# Estimating the reproduction number of COVID-19 in Iran using epidemic modeling

**DOI:** 10.1101/2020.03.20.20038422

**Authors:** Ebrahim Sahafizadeh, Samaneh Sartoli

**Affiliations:** Department of Information Technology, Payame Noor University, P, O, Box 19395-3697 Tehran, IRAN

**Keywords:** COVID-19, coronavirus, Iran, basic reproduction number, epidemiolgy, SIR

## Abstract

**Background:** As reported by Iranian governments, the first cases of coronavirus (COVID-19) infections confirmed in Qom, Iran on February 19, 2020 (30 Bahman 1398). The number of identified cases afterward increased rapidly and the novel coronavirus spread to all provinces of the country. This study aimed to fit an epidemic model to the reported cases data to estimate the basic reproduction number (*R*_0_) of COVID-19 in Iran.

**Methods:** We used data from February 21, 2020, to April 21, 2020, on the number of cases reported by Iranian governments and we employed the SIR (Susceptible-Infectious-Removed) epidemic spreading model to fit the transmission model to the reported cases data by tuning the parameters in order to estimate the basic reproduction number of COVID-19 in Iran.

**Results:** The value of reproduction number was estimated 4.86 in the first week and 4.5 in the second week. it decreased from 4.29 to 2.37 in the next four weeks. At the seventh week of the outbreak the reproduction number was reduced below one.

**Conclusions:** The results indicate that the basic reproduction number of COVID-19 was significantly larger than one in the early stages of the outbreak. However, implementing social distancing and preventing travelling on Nowruz (Persian New Year) effectively reduced the reproduction number. Although the results indicate that reproduction number is below one, it is necessary to continue social distancing and control travelling to prevent causing a second wave of outbreak.

## Background

On December 31, 2019, the novel coronavirus (COVID-19) emerged in Wuhan, China [1] and the potential of transmitting virus via human travelling caused to spread the novel corona virus to other Chinese provinces as well as the other countries [2].

Iranian governments reported the first confirmed cases of novel coronavirus (COVID-19) infections in Qom, on February 19, 2020 (30 Bahman 1398) [3].

Tehran, Guilan and Markazi were the other provinces confirmed to be infected by coronavirus during the next days of the outbreak. After one week Iranian governments confirmed that 18 out of 31 provinces of Iran were infected by the novel coronavirus. After two weeks the number of reported cases increased to 2922 people of whom 92 cases died and 552 cases recovered [3].

The aim of this study is to estimate basic reproduction number (R0) of COVID-19 in Iran. For this purpose, we use the reported data by Iranian governments and develop an epidemic model to fit on the real data by tuning the model parameters. We then use the model parameters’ values to calculate the basic reproduction number of COVID-19 in Iran.

## Methods

We employ the SIR (Susceptible-Infected-Removed) epidemic model to represent the spreading process of COVID-19 in Iran. SIR model is an epidemic spreading model proposed by Kermack and McKendric [4]. We estimate the parameters of model to get a best fit on reported data of COVID-19 outbreak in Iran. The differential equations of the SIR model are given as [5]:

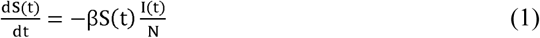

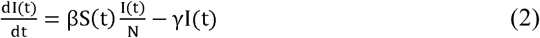

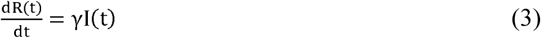

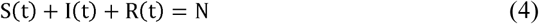

Where *S(t), I(t)* and *R(t)* are the number of susceptible, infected and removed people. We assume that removed cases are the summation of recovered and dead cases. *β* is the infection rate, and *γ* is the remove rate which is the inverse of infectious period.

The basic reproduction number, *R*_0_, is the average number of the secondary individuals in a complete susceptible population infected by a single infected person during its spreading life [6]. When *R*_0_ > 1 virus spreads through the population and when *R*_0_ < 1 the outbreak will stop due to decreasing the number of new cases. According to Eq. (2) when the number of infected people increases we have:

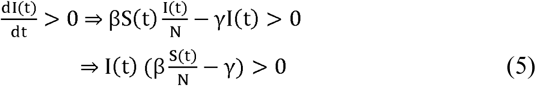

In a complete susceptible population which S(t) ≈ N the Eq. (5) can be written as the following:

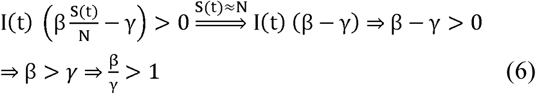

According to Eq. 6 and definition of *R*_0_ we have:

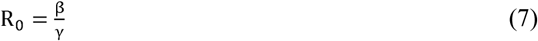

Due to the infection of all provinces of Iran, N is assumed to be the population of Iran (81800269 in 2018) which was obtained from the World Bank [7].

We model epidemic spreading in the period from February 21, 2020 to April 21, 2020. Table 1. Shows the number of reported infected cases and reported removed cases by Iran ministry of health and medical education (MOHME) [3]. MOHME reported the first two cases of COVID-19 infections and their death on 19 February, 2020 which is indicated as day zero in Table 1.

**Table 1.** The number of infectious and removed people reported by Iranian governments

| Date | Day | Number of infected cases | Number of removed cases (recovered+dead) | Date | Day | Number of infected cases | Number of removed cases (recovered+dead) |
| --- | --- | --- | --- | --- | --- | --- | --- |
| 19/02/2020 | 0 | 0 | 2 | 22/03/2020 | 32 | 12040 | 9598 |
| 20/02/2020 | 1 | 3 | 2 | 23/03/2020 | 33 | 12861 | 10188 |
| 21/02/2020 | 2 | 14 | 4 | 24/03/2020 | 34 | 13964 | 10847 |
| 22/02/2020 | 3 | 23 | 5 | 25/03/2020 | 35 | 15315 | 11702 |
| 23/02/2020 | 4 | 35 | 8 | 26/03/2020 | 36 | 16715 | 12691 |
| 24/02/2020 | 5 | 46 | 15 | 27/03/2020 | 37 | 18821 | 13511 |
| 25/02/2020 | 6 | 77 | 18 | 28/03/2020 | 38 | 21212 | 14196 |
| 26/02/2020 | 7 | 96 | 43 | 29/03/2020 | 39 | 23278 | 15031 |
| 27/02/2020 | 8 | 167 | 80 | 30/03/2020 | 40 | 24827 | 16668 |
| 28/02/2020 | 9 | 281 | 107 | 31/03/2020 | 41 | 27052 | 17554 |
| 29/02/2020 | 10 | 427 | 166 | 01/04/2020 | 42 | 29084 | 18509 |
| 01/03/2020 | 11 | 749 | 229 | 02/04/2020 | 43 | 30592 | 19871 |
| 02/03/2020 | 12 | 1144 | 357 | 03/04/2020 | 44 | 31936 | 21247 |
| 03/03/2020 | 13 | 1824 | 512 | 04/04/2020 | 45 | 32555 | 23188 |
| 04/03/2020 | 14 | 2278 | 644 | 05/04/2020 | 46 | 32612 | 25614 |
| 05/03/2020 | 15 | 2667 | 846 | 06/04/2020 | 47 | 32525 | 27975 |
| 06/03/2020 | 16 | 3710 | 1037 | 07/04/2020 | 48 | 31678 | 30911 |
| 07/03/2020 | 17 | 4009 | 1814 | 08/04/2020 | 49 | 30781 | 33805 |
| 08/03/2020 | 18 | 4238 | 2328 | 09/04/2020 | 50 | 29801 | 36419 |
| 09/03/2020 | 19 | 4530 | 2631 | 10/04/2020 | 51 | 28495 | 39697 |
| 10/03/2020 | 20 | 5020 | 3022 | 11/04/2020 | 52 | 23698 | 46331 |
| 11/03/2020 | 21 | 5687 | 3319 | 12/04/2020 | 53 | 23318 | 48368 |
| 12/03/2020 | 22 | 6370 | 3705 | 13/04/2020 | 54 | 22735 | 50568 |
| 13/03/2020 | 23 | 7321 | 4043 | 14/04/2020 | 55 | 22065 | 52812 |
| 14/03/2020 | 24 | 7779 | 4950 | 15/04/2020 | 56 | 21679 | 54710 |
| 15/03/2020 | 25 | 8624 | 5674 | 16/04/2020 | 57 | 20897 | 57098 |
| 16/03/2020 | 26 | 9142 | 5849 | 17/04/2020 | 58 | 20472 | 59022 |
| 17/03/2020 | 27 | 9792 | 6377 | 18/04/2020 | 59 | 19850 | 61018 |
| 18/03/2020 | 28 | 10516 | 6845 | 19/04/2020 | 60 | 20070 | 62141 |
| 19/03/2020 | 29 | 11144 | 7263 | 20/04/2020 | 61 | 19023 | 64482 |
| 20/03/2020 | 30 | 11466 | 8178 | 21/04/2020 | 62 | 18540 | 66262 |
| 21/03/2020 | 31 | 11119 | 9191 |  |  |  |  |

## Results

The Runge-Kutta method is used to resolve the SIR model’s ODEs to fit model on reported data. Numerical simulation is conducted in MATLAB and we tune the parameters β and γ to have the best fit plot on reported data plot. We then estimate the basic reproduction number by dividing *β* on *γ*. The SIR model fitting results are shown in Fig. 1 Through Fig. 4. The red and green plots are model plots and the blue and pink plots are reported data plots. As shown in Fig. 1, we estimated that the *R*_0_ to be 4.86 in the first week of outbreak which is significantly larger than one. We then set *β* = 0.585 and *γ* = 0.13 to fit model to reported data of the second week and we obtained *R*_*t*_ = 4.5 which is smaller than the first week. In Fig. 3 we obtained R_t_ = 4.29 for the 13^th^ day to 16^th^ day of outbreak. Fig. 4 shows that the reproduction number significantly decreased to 2.37 between March 6, 2020 to April 3, 2020, which could be the consequence of self-quarantining and reducing working time by government. As Nowruz occurred on March 20, 2020, many businesses as well as government offices closed and the government restricted travelling between provinces for almost three weeks. As a consequence of this policy it can be seen at Fig 5. that the reproduction number declined to one between April 4 and April 6, 2020. Fig. 6 shows that since April 7, 2020 the reproduction number has been less than one which indicates that disease will decline.

**Fig. 1:**
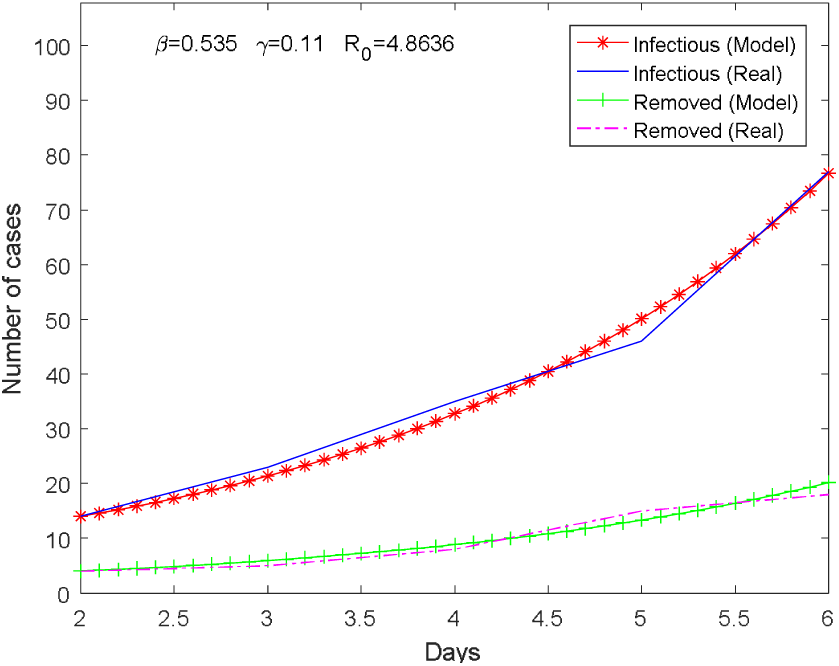
Plot of COVID-19 outbreak on the first week

**Fig. 2:**
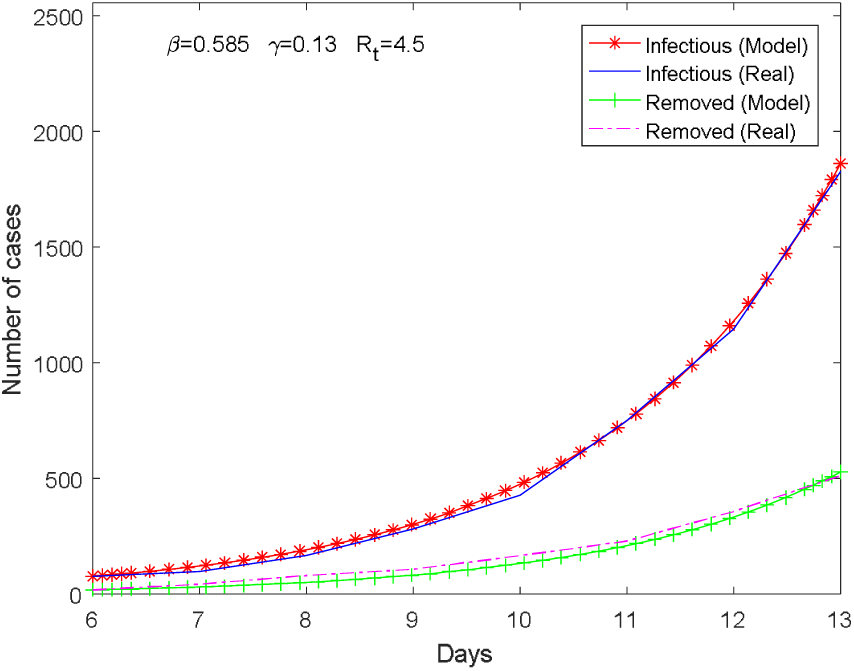
Plot of COVID-19 outbreak on the second week

**Fig. 3:**
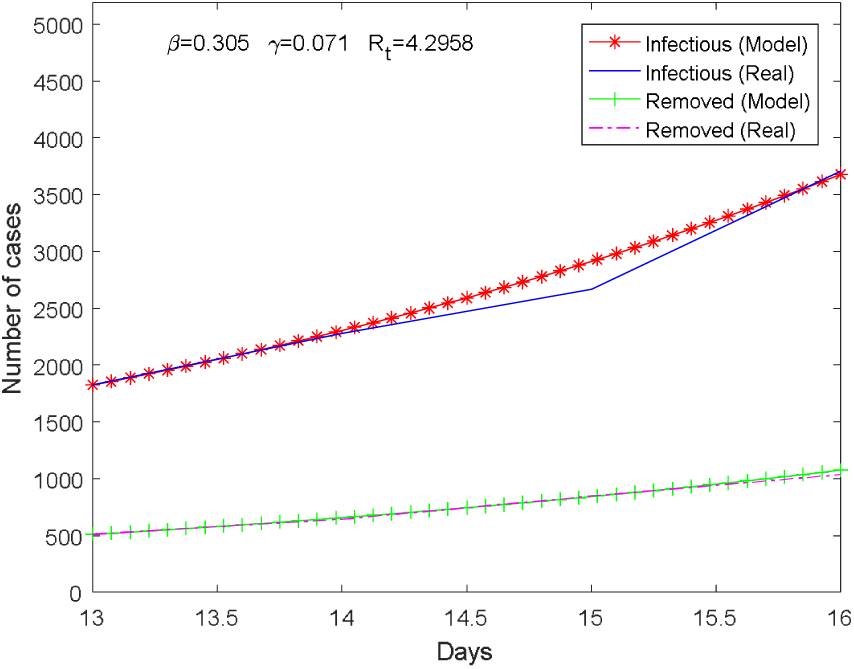
Plot of COVID-19 outbreak from the 13^th^ day to 16^th^.

**Fig. 4:**
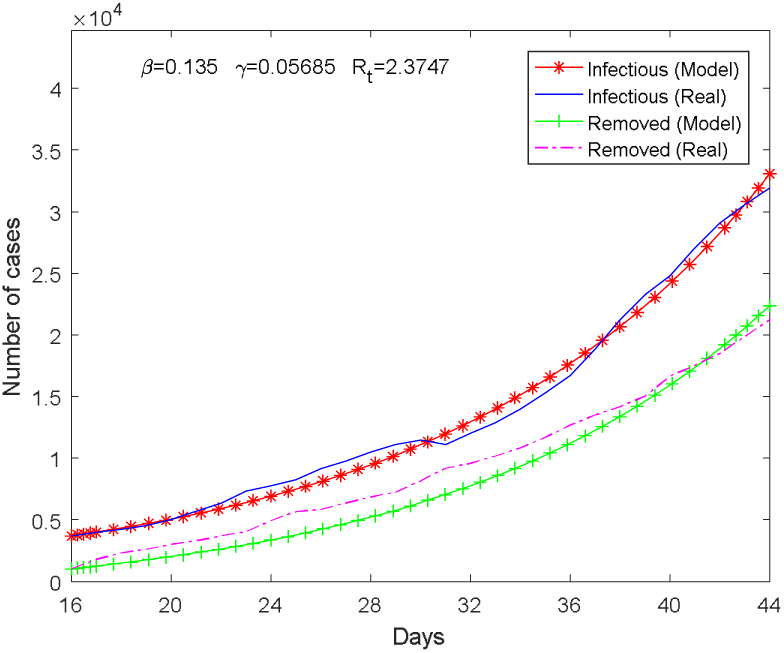
Plot of COVID-19 outbreak from the 16^th^ day to 44^th^.

**Fig. 5:**
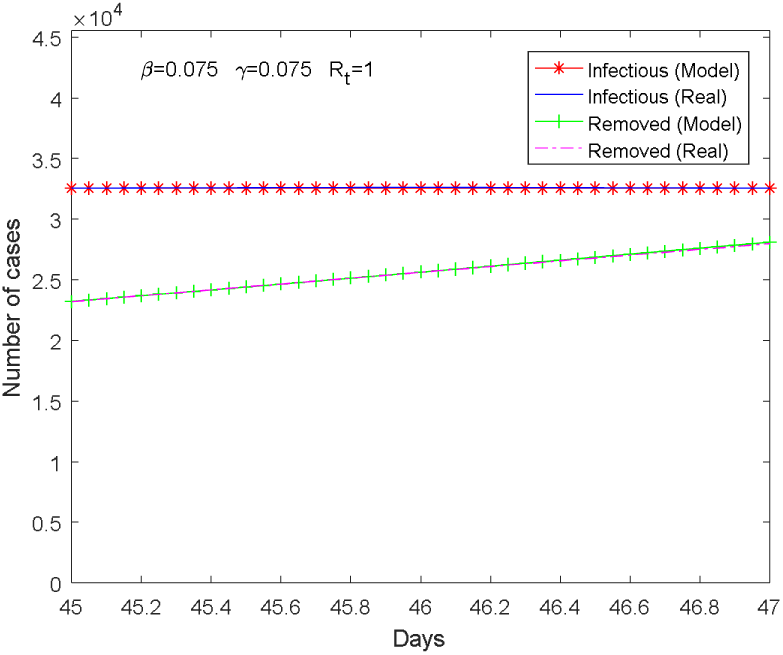
Plot of COVID-19 outbreak from the 45^th^ day to 47^th^.

**Fig. 6:**
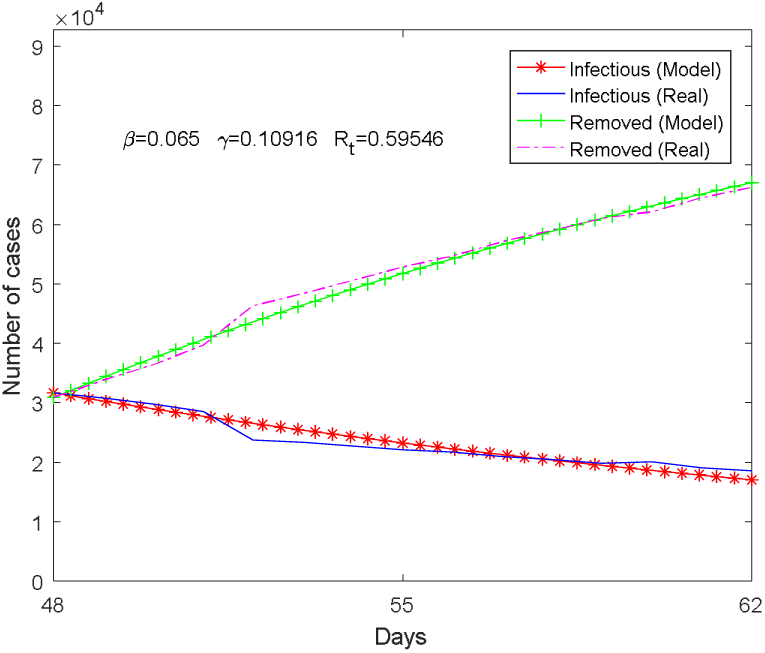
Plot of COVID-19 outbreak from the 48^th^ day to 62^th^.

## Discussion

In this study, we employed SIR epidemic model, to fit the reported data in Iran and estimate the basic reproduction number of COVID-19 in Iran.

Previous studies estimated basic reproduction number between 2.2 and 4. Chen et.al [1] showed that the basic reproduction number of COVID-19 was 3.58 using the reported data in Wuhan City, China. Read et.al [8] showed that the *R*_0_ of COVID-19 was between 3.6 and 4. Zhao et.al [9] estimated *R*_0_ of COVID-19 to be between 2.24 and 3.58. Other researchers estimated *R*_0_ to be 2.68 [10], 2.6 [2] and 2.2 [11].

The simulation results of this study indicated that the *R*_0_ of COVID-19 was 4.86 in the first week of the outbreak which was significantly larger than one. The schools and universities closed in the first week and in the second week the effective reproduction number decreased to 4.5. In the second week the working time reduced and effective reproduction number decreased to 4.29 from 13^th^ day to 16^th^. Comparing to the other researches, the results showed that R_0_ of COVID-19 in Iran in the early stages were higher than the previous estimated basic reproduction number [1][2][8][9][10][11].

In the next weeks of outbreak people decided to stay at home spontaneously and the government reduced the working time. The effective reproduction number decreased to 2.37 between March 6, 2020 and April 3, 2020. In Nowruz, the government restricted travelling and many businesses closed for three weeks. Hence, the reproduction number was reduced and it has been less than one since April 7.

## Conclusion

Basic reproduction number of COVID-19 is an important parameter which is used to estimate the risk of COVID-19 outbreak. We estimated the basic reproduction number (R_0_) of COVID-19 outbreak in Iran using SIR model by fitting the model to official reported data. According to the results, it is observed that reproduction number of COVID-19 in Iran in the early stages 4.86, 4.5 and 4.29 are higher than the previous estimated [1][2][10].

Self-quarantine and control strategies implemented by government reduced the effective reproduction number of COVID-19 below one. However, it is necessary to continue social distancing and control travelling to prevent causing a second wave of outbreak.

## Data Availability

Information related to the study is in the manuscript

## Abbreviations

Covid-19 coronavirus; basic reproduction number; Iran.

## Authors’ contributions

ES conducted the experiments and analyzed the results and wrote the manuscript; SS collected the reported cases data and review and edited the manuscript. All authors read and approved the final manuscript.

## Funding

The authors received no specific funding for this work.

## Availability of data and materials

Information related to the study is in the manuscript.

## Ethics approval and consent to participate

Not applicable.

## Consent for publication

Not applicable.

## Competing interests

The authors declare that they have no competing interests.

## Notes

### Competing Interest Statement

The authors have declared no competing interest.

